# Seroepidemiology of COVID-19 in pregnant women and their infants in Uganda and Malawi across multiple waves 2020-2022

**DOI:** 10.1101/2023.08.19.23294311

**Authors:** Lauren Hookham, Liberty Cantrell, Stephen Cose, Bridget Freyne, Luis Gadama, Esther Imede, Kondwani Kawaza, Samantha Lissauer, Phillipa Musoke, Vicki Nankabirwa, Musa Sekikubo, Halvor Sommerfelt, Merryn Voysey, The periCOVID Consortium, Kirsty Le Doare

**Author notes:** Membership of the periCOVID Consortium is provided in the Acknowledgments.

## Abstract

Data on SARS-CoV-2 infection in pregnancy and infancy has accumulated throughout the course of the pandemic. However, limited information is available from countries in sub-Saharan Africa (SSA). Evidence regarding asymptomatic SARS-CoV-2 infection and adverse birth outcomes are also scarce in these countries. The pregnant woman and infant COVID in Africa study (PeriCOVID Africa) is a South-South-North partnership involving hospitals and health centres in five countries: Malawi, Uganda, Mozambique, The Gambia, and Kenya. The study leveraged data from three ongoing prospective cohort studies: Preparing for Group B Streptococcal Vaccines (GBS PREPARE), SARS-CoV-2 infection and COVID-19 in women and their infants in Kampala and Mukono (COMAC) and Pregnancy Care Integrating Translational Science Everywhere (PRECISE). In this paper we describe the seroepidemiology of SARS-CoV-2 infection in pregnant women enrolled in sites in Uganda and Malawi, and the impact of SARS-CoV-2 infection on pregnancy and infant outcomes.

The PeriCOVID study is a prospective mother-infant cohort study that recruited pregnant women at any gestation antenatally or on the day of delivery. A nasopharyngeal swab was taken from mothers at enrolment for RT-PCR confirmation of SARS-CoV-2 infection, and maternal and cord blood samples were tested for SARS-CoV-2 antibodies using Wantai and Euroimmune ELISA. The primary outcome was seroprevalence of SARS-CoV-2 antibodies in maternal blood, reported as the proportion of seropositive women by study site and wave of COVID-19 within each country. Placental transfer of antibodies was described using the geometric mean ratio (GMR). We also estimated the proportion of asymptomatic or subclinical COVID-19 infections in pregnant women using serological testing and collected adverse pregnancy and infancy outcomes (e.g. still-birth, prematurity, maternal or infant death).

In total, 1379 women were enrolled, giving birth to 1387 infants. Overall, 63% of pregnant women had a SARS-CoV-2 positive serology. Over subsequent waves (delta and omicron), in the absence of vaccination, seropositivity rose from 20% to over 80%. The placental transfer GMR was 1.7, indicating active placental transfer of anti-spike IgG. There was no association between SARS-CoV-2 antibody positivity and adverse pregnancy or infancy outcomes. This study describes the increasing prevalence of SARS CoV-2 antibodies in pregnant woman in Uganda and Malawi across waves of SARS-CoV-2 infection. Our study adds to existing evidence that suggests under-reporting of infection if based solely on cases with clinical disease, or a positive RT-PCR for SARS-CoV-2, as most of the women in our study had asymptomatic infections and did not seek medical care. This has implications for screening in subsequent outbreaks and pandemics where protection of pregnant women and effect of infection in pregnancy on the infant are unknown.

## Introduction

The initial predictions of the impact of SARS-CoV-2 in sub-Saharan Africa suggested high case numbers and fatalities (1), yet there is evidence to suggest that the pandemic evolved differently in Africa than in other regions (2, 3). Several factors have been proposed to explain the relatively low frequency of severe SARS-CoV-2 illness, including a younger population, a lack of long-term care facilities and reduced population density (4–7). However, limited testing capacity and weak reporting structures in many sub-Saharan African countries may also result in under-reporting, leading to an underestimation of the true risk of serious SARS-CoV-2 infection (3). Infection in pregnancy, even if asymptomatic or mild, may have long-term impacts for a pregnant woman or impair the neurodevelopment of her child, a risk which is well established for viral infections such as Zika virus or cytomegalovirus (8).Data on the impact of SARS-CoV-2 infection in pregnancy on neurodevelopmental outcomes is emerging (9, 10). Evidence regarding asymptomatic SARS-CoV-2 infection and adverse birth outcomes is limited and the long-term effects of the pandemic on infant health remain poorly understood (11).

Serological surveillance is a useful means of estimating population-level immunity against infectious diseases using cross-sectional studies of antibody prevalence (12). In the case of SARS-CoV-2, serological surveys are helpful in estimating the number of people who have been exposed to SARS-CoV-2, whether they were symptomatic or not to better clarify the dynamics of exposure during the different epidemic waves as vaccines were rolled out (13). For example, seroprevalence surveys conducted across Kenya, South Africa and Malawi have all reported community transmission, which is several times higher than that detected by national virological surveillance programmes (14–16).

Seroprevalence studies in pregnancy enables insights into the real magnitude of exposure to SARS-CoV-2 infections and the extent of under-reporting of SARS-CoV-2 cases. Information on seroprevalence in pregnancy, placental antibody transfer and antibody half-life also offer the possibility to approximate the number of mother-infant dyads who could potentially exhibit immunological protection against subsequent infections, especially with low vaccine coverage in many low-resource settings (LRS). Finally, seroprevalence studies can also provide insight into the relationship between infection and vaccination, symptoms, and antibody responses to assist with future screening and prevention policies in pregnancy.

To address these specific gaps, we investigated the seroprevalence and the associations of different factors on seropositivity to the SARS-CoV-2 virus among pregnant women and their infants in Uganda and Malawi. This was performed during consecutive SARS-CoV-2 waves.

## Methods

### Study design and participants

PeriCOVID Africa is a multi-site prospective mother-infant cohort study using an adapted WHO UNITY protocol (17), whereby women were categorised into two categories dependent on serological testing using the Wantai total antibody assay as exposed (positive serology) or unexposed (negative serology) to SARS-CoV-2. Women were additionally screened for symptoms, using the WHO definition for probable COVID-19 disease at the time of study participation (18). We defined symptomatic COVID-19 infection according to the WHO definitions of probable COVID-19 illness (18) and asymptomatic infection as seropositive or PCR positive at enrolment in the absence of reported symptoms. Unexposed women were those with no reported symptoms consistent with SARS-CoV-2 infection and negative serology.

#### Recruitment

Each study adapted the WHO UNITY protocol according to local needs and capacity considerations. Women were recruited into the study either during an antenatal visit, or during labour at seven study clinics and hospitals in Uganda and Malawi. In all studies, gestational age at enrolment was estimated by date of last menstrual period and fundal height. Additionally, in periCOVID Uganda and periCOVID Malawi Ballard scores were calculated at birth. Individual study recruitment, sampling and follow up are shown in Table 1. The first participant was recruited 1^st^ February 2021. Final participant follow and sampling was concluded by 31^st^ January 2022.

**Table 1.** Recruitment, sampling and follow-up by study site.

| Study Site | Enrolment timing | Study period | Inclusion Criteria | Exclusion Criteria | Sampling at enrolment | Follow up |
| --- | --- | --- | --- | --- | --- | --- |
| All sites |  |  | Willingness to provide informed consent |  | Maternal and Cord blood | 6 week follow up |
| periCOVID Uganda | Antenatal clinic or delivery | February 2021 – January 2022 | Pregnant women (including emancipated minors aged over 14 years) at any gestation including the day of delivery<br>Planning to deliver at one of the designated study sites and willing to stay in the area for the first six weeks of their baby's life<br>Willing to attend a follow up visit at six weeks postpartum | No exclusion criteria | Maternal nasopharyngeal swab* | **For COVID-19 Cases only\*. Maternal:** nasopharyngeal maternal blood sample (5ml serum), **Infant:** nasal swab, blood sample (2-5ml) or dried blood spot, |
| COMAC | Delivery | August 2021 – January 2022 | - low risk of infection with tuberculosis in the household<br>-mother of legal age (including if emancipated minor) for participation<br>-mother residing within the study area, not intending to move out | - Baby weighs less than 2kg at birth<br>- Baby requires hospital admission for severe illness at birth<br>- Serious congenital malformation(s)<br>- severely ill mother on the day of giving birth whose condition(s) | Nil additional | **Maternal** blood test at 14 weeks. Nasopharyngeal swab at 6 weeks and 14 weeks. **Infant:** blood test at 14 weeks. Nasopharyngeal swab at 6 weeks and 14 weeks |
|  |  |  | of the area in the next 4 months and is likely to be traceable for up to 12 months.<br>- HIV-1 positive women should be receiving the necessary antiretroviral treatment and prophylaxis (Ugandan Option B+ guidelines)<br>HIV-exposed babies receives peri exposure prophylaxis (Ugandan Option B+ guidelines) | require(s) hospitalization<br>- |  |  |
| Malawi | Delivery | March 2021 – January 2022 | All women presenting to QECH in labour with an estimated gestation of 28 weeks or greater who<br>- Are willing to attend a follow up visit at 6 weeks postpartum | No exclusion criteria. | Nil additional | **For COVID-19 cases only. Maternal**<br>*blood, rectal swab, breastmilk. **Infant\*:**<br>blood |

#### COVID-19 testing

All women at all study sites were screened for COVID-19 symptoms using a standardised data collection form with questions including a recent history of fever, cough, anosmia and ageusia and contact with a known SARS-Cov-2 case. Information on COVID-19 illness symptoms was collected at enrolment for the 14 days prior to enrolment in PeriCOVID Uganda and Malawi, and in the 28 days prior to enrolment in COMAC Uganda. Women enrolled in PeriCOVID Uganda had a nasal swab taken at enrolment to test for SARS-CoV-2 by PCR. In Malawi throat swabs were taken if the clinical syndrome was suggestive of a probable COVID-19 illness as defined by the WHO (18).

#### Blood sampling

Sampling at all sites for antibodies to SARS-COV-2 included at least a maternal venous and paired cord blood sample (see Table 1 for sampling schedule at each site).

#### Data collection

Each site (KNRH, Kawaala, Kitebi or Mukono General Hospital in Kampala, Uganda and QECH in Blantyre, Malawi) used a study questionnaire which was completed by research staff to capture information from study participants and then uploaded to a central RedCAP database. This included data on maternal age, significant past medical history, HIV and socioeconomic status; onset and duration of signs and symptoms of SARS-CoV-2 illness if present, and self-reporting of prior SARS-CoV-2 illness; gestational age at enrolment, parity, number of foetuses (if known before delivery), co-infection with malaria, vaccinations received in pregnancy including the SARS-CoV-2 vaccines; gestational age at delivery, delivery method, intrapartum and postpartum complications such as pre-term birth, stillbirth, abortion, and maternal death; neonatal outcomes including evidence of COVID-19 illness, Neonatal Intensive Care Unit (NICU) admission, low birth weight and neonatal death; infant health status

### Laboratory methods

As per FIND guidelines at the time of the study protocol development (19), we used three different SARS-CoV-2 specific antibody assays that targeted either the receptor binding domain (RBD; total antibody), the spike protein (anti-S, total antibody) or nuclear capsid (anti-NCP, IgG). We performed in-house specificity and sensitivity testing, respectively, using 100 pre-COVID19 (pre-2019) samples selected by month for seasonality assessment and 20 PCR positive samples (19) to perform assay validation. We also examined potential cross-reactivity in our assay from malaria-specific antibodies using 74 women who tested positive to malaria (antibody positive by rapid diagnostic test (RDT) from pre-COVID samples and 15 SARS-COV-2 PCR positive samples in women who did not have malaria (negative RDT) during the COVID-19 pandemic. Results can be seen in Supplementary Table S1.

#### Laboratory analysis

Laboratory testing for SARS-COV-2 antibodies was performed at the MRC/UVRI and LSHTM Uganda Research Unit or the Malawi Liverpool Wellcome facilities using the Wantai SARS-CoV-2 total antibody ELISA kit (Beijing Wantai Biological Pharmacy Enterprise Co., Ltd, Beijing, China). The manufacturer-reported assay sensitivity is 94.4%, with a specificity of 100%. All specimens that tested positive for Wantai were tested using Euroimmun Anti-SARS-CoV-2 NCP/S ELISA (IgG) (Euroimmun, Lübeck, Germany) kits for the detection of IgG antibodies to SARS-COV-2 nucleocapsid and spike (S1 and S2) proteins, respectively. Euroimmun Anti-SARS-CoV-2 NCP/S ELISA (IgG) is a semi-quantitative immunoassay with a reported sensitivity of 94.6% and specificity of 99.8% in samples collected at least 10 days after confirmed SARS-COV-2 infection. A sample was considered positive if the Wantai test was positive. Results which were reported as borderline on the Euroinmmune assay were considered as negative for the purpose of our analysis. Due to variable specificity of the Euroimmune assay, we report Wantai results for all outcomes as per manufacturer’s instructions.

As the Wantai ELISA is a qualitative test, WHO standards for NCP and S proteins were run on all assays. The stock concentration for NCP and S proteins was 123µg/ml and 1000µg/ml, respectively. The working concentration for both NCP and S proteins was 2 µg/ml. The calibration curve was created using WHO International standard for anti-SARS-CoV-2 immunoglobulin (NIBSC 20/136) using a 12-well dilution series created in 1.75-fold steps, starting at 1:200 and this series was used to generate the curve. All laboratory testing in Uganda for SARS-CoV-2 antibodies was performed using the ETI-MAX 3000 (Diasorin, Saluggia, Italy).

### Data Analysis

Although there was no predefined sample size, we aimed to obtain samples from at least 1000 women and infants across all sites. The sero-epidemiological analyses were carried out using all participants with enrolment blood sample results available for analysis. The proportion of seropositive results was calculated for the individual waves of SARS-CoV-2 within each country. This was done for maternal blood samples to estimate the seroprevalence of SARS-CoV-2 among pregnant women, and for cord blood samples to estimate the seroprevalence of SARS-CoV-2 antibodies in infants. The dates used to define the SARS-COV-2 waves in each country are given in Table S2 and were taken from Our World in Data (20). Waves were defined by taking the nadir between each peak for the start and end dates of each wave. Confidence intervals for prevalence estimates were computed using the Clopper-Pearson (Binomial Exact) method.

The geometric mean concentration (GMC) and 95% confidence intervals (CI) of anti-S and anti-NCP antibodies measured on the Euroimmune assay were calculated for mother-infant pairs. To study the rate of placental transfer of SARS-CoV-2 antibodies, the geometric mean ratio (GMR) of infant to maternal antibodies was calculated.

The proportion of participants with a symptomatic or asymptomatic infection was calculated for women who were seropositive at enrolment and for those who had a positive RT-PCR test at enrolment.

The impact of infection on key pregnancy and neonatal outcomes was modelled for women and infants in PeriCOVID Malawi and Uganda log-binomial generalised linear models (GLM) which were adjusted for country. Models were constructed for maternal death, infant death, combined adverse pregnancy outcome (at least one of maternal death, abortion, premature labour or stillbirth), and the combined adverse neonatal outcome (at least one of neonatal/infant death, prematurity, low birth weight, NICU admission after birth, or birth asphyxia). Relative risks were presented with 95% confidence intervals for all models reaching convergence. Results were not presented for adverse outcomes with fewer than 5 events.

Statistical analyses were carried out using R version 4.2.1. No significance tests were conducted.

### Ethical Considerations

The study documents were reviewed and approved by the Ethics Committee of the relevant institutions: Uganda: Makarere University School of Medicine (SOMREC), Uganda National Council for Science and Technology (UNCST); Regional Committees for Medical and Research Ethics in Norway; Malawi: College of Medicine Research Ethics Committee (COMREC).

## Results

In total, 1379 women were enrolled, giving birth to 1387 infants (Fig 1, Table 2). The mean age of all women was 26 years (SD 6) across the three sites. Most (n=1346, 98%) pregnancies were singleton. The HIV prevalence was 9% (n= 272). 371 women delivered outside of a study hospital and so no blood samples were available for analysis. Deliveries of 1024 infants with cord blood samples occurred at study sites. Almost all were livebirths (n=1009/1024, 99%) and most (n=888/1024, 87%) were not admitted to the NICU after birth (Table 2). A total of 909/1379 (65.9%) women in periCOVID Uganda and periCOVID Malawi had a PCR result at enrolment available for analysis of which 68/909 (7.5%) women had positive PCR results for SARS-CoV-2. Amongst these women, 77.9% (n=53) had symptoms consistent with COVID-19 disease. The majority (88.7%, n=47/53) were in Malawi who were performing RT-PCR testing only on symptomatic women. In periCOVID Uganda, where all women had a RT-PCR test at enrolment, 31.6% (n=6/19) of those with a positive RT-PCR test had symptoms suggestive of COVID-19 disease (Supplemental table 3). Genotyping revealed all positive cases from Uganda to be of the delta variant.

**Fig 1.**
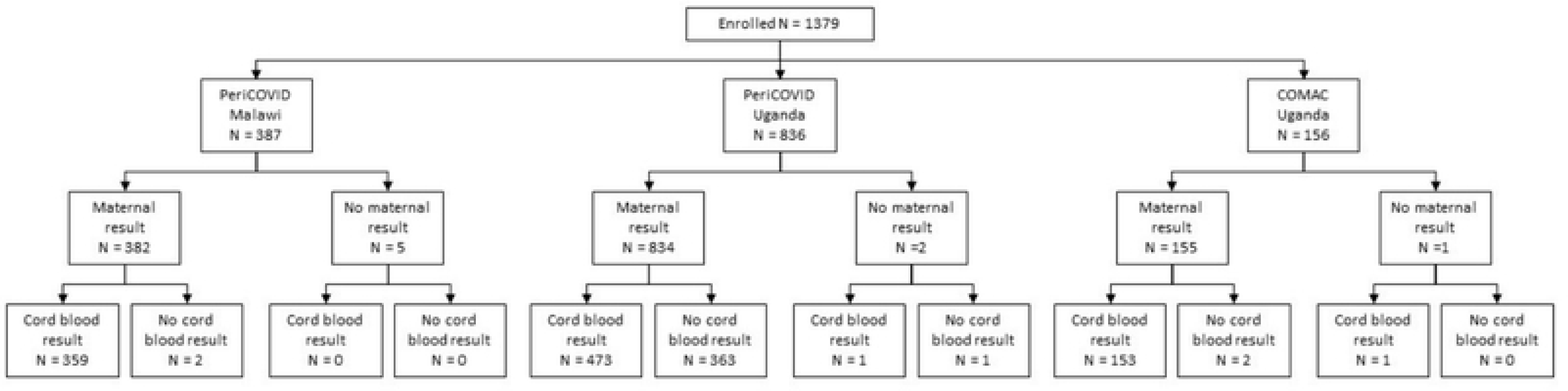
Study flow chart. Study flow chart to show the number of women enrolled by study, with maternal serology available at enrolment (maternal result) and the infants with cord blood serology (cord blood result) available.

**Table 2.** Demographics for women and infants in the study.

| Characteristic of women enrolled | Overall, N = 1,379 | PeriCOVID Malawi, N = 387 | PeriCOVID Uganda, N = 836 | COMAC Uganda, N = 156 |
| --- | --- | --- | --- | --- |
| Age (years) | 26 (6) | 25 (7) | 26 (6) | 27 (5) |
| Missing | 2 | 2 | 0 | 0 |
| Number of fetuses |  |  |  |  |
| Singleton | 1,346 (98%) | 371 (96%) | 819 (98%) | 156 (100%) |
| Twins | 30 (2.2%) | 14 (3.6%) | 16 (1.9%) | 0 (0%) |
| Triplets | 1 (<0.1%) | 0 (0%) | 1 (0.1%) | 0 (0%) |
| Missing | 2 | 2 | 0 | 0 |
| Highest level of education |  |  |  |  |
| Graduate Education /Terminal Degree Completed. | 23 (1.7%) | 1 (0.3%) | 20 (2.4%) | 2 (1.3%) |
| No Formal Education | 12 (0.9%) | 5 (1.3%) | 5 (0.6%) | 2 (1.3%) |
| Other | 2 (0.1%) | 0 (0%) | 2 (0.2%) | 0 (0%) |
| Primary Education Completed | 211 (15%) | 98 (26%) | 84 (10%) | 29 (19%) |
| Secondary Education Completed | 252 (18%) | 129 (34%) | 105 (13%) | 18 (12%) |
| Some Primary Education | 239 (17%) | 126 (33%) | 86 (10%) | 27 (17%) |
| Some Secondary Education | 509 (37%) | 0 (0%) | 432 (52%) | 77 (49%) |
| University / College Completed | 127 (9.2%) | 25 (6.5%) | 101 (12%) | 1 (0.6%) |
| Missing | 4 | 3 | 1 | 0 |
| HIV |  |  |  |  |
| No | 1,107 (91%) | 325 (84%) | 782 (94%) | 0 (NA%) |
| Yes | 114 (9.3%) | 60 (16%) | 54 (6.5%) | 0 (NA%) |
| Missing | 158 | 2 | 0 | 156 |
| **Characteristics of infants born and providing a cord blood sample** | **Overall, N = 1,024** | **PeriCOVID Malawi, N = 365** | **PeriCOVID Uganda, N = 503** | **COMAC Uganda, N = 156** |
| Status of infant at birth |  |  |  |  |
| Livebirth | 1,009 (99%) | 357 (99%) | 496 (99%) | 156 (100%)* |
| Miscarriage | 2 (0.2%) | 1 (0.3%) | 1 (0.2%) | 0 (0%) |
| Stillbirth | 10 (1.0%) | 4 (1.1%) | 6 (1.2%) | 0 (0%) |
| Missing | 3 | 3 | 0 | 0 |
| Gestation at birth (weeks) | 27, 44 | 27, 44 | 28, 44 | Inf, -Inf |
| Missing | 157 | 1 | 0 | 156 |
| Sex |  |  |  |  |
| Female | 492 (48%) | 164 (45%) | 248 (49%) | 80 (51%) |
| Male | 531 (52%) | 200 (55%) | 255 (51%) | 76 (49%) |
| Missing | 1 | 1 | 0 | 0 |
| Birth weight (grams) |  |  |  |  |
| Mean (SD) | 3,082 (540) | 2,910 (573) | 3,151 (504) | 3,262 (458) |
| Range | 600, 4,850 | 600, 4,500 | 1,200, 4,850 | 2,100, 4,700 |
| Missing | 8 | 2 | 6 | 0 |
| Admitted to NICU after birth |  |  |  |  |
| No | 888 (87%) | 264 (73%) | 468 (93%) | 156 (100%)* |
| Yes | 133 (13%) | 98 (27%) | 35 (7.0%) | 0 (0%) |
| Missing | 3 | 3 | 0 | 0 |

259 \* COMAC Uganda only recruited infants who were born alive and not admitted to the NICU after birth

### Seropositivity in pregnant women and their infants

Overall, 1371/1379 maternal samples (382 from Malawi and 989 from Uganda) and 987/1024 cord blood samples (359 from Malawi and 628 from Uganda) were available for analysis (Table 3 and 4). There were 875 SARS-CoV-2 seropositive women in the study (257 in Malawi, 618 in Uganda), of whom 791 (90.4%) were asymptomatic in the 14 days (periCOVID Uganda and Malawi) −28 days (COMAC) prior to enrolment. This corresponded in Malawi to 21 asymptomatic seropositive participants during the second wave, 134 during the third wave and 57 in the fourth wave. In Uganda, this corresponded to 92 in the first wave, 422 in the second wave and 65 in the third wave (Table 3).

There was an increase in seropositivity with each subsequent wave, and the majority (70-100%) of women with positive serology were asymptomatic in the 14 days prior to sampling. Fig 2 shows the monthly total antibody positivity rate for each site with the daily number of new cases per million in Uganda and Malawi. The concordance between maternal and cord samples was high (72-99%).

**Fig 2.**
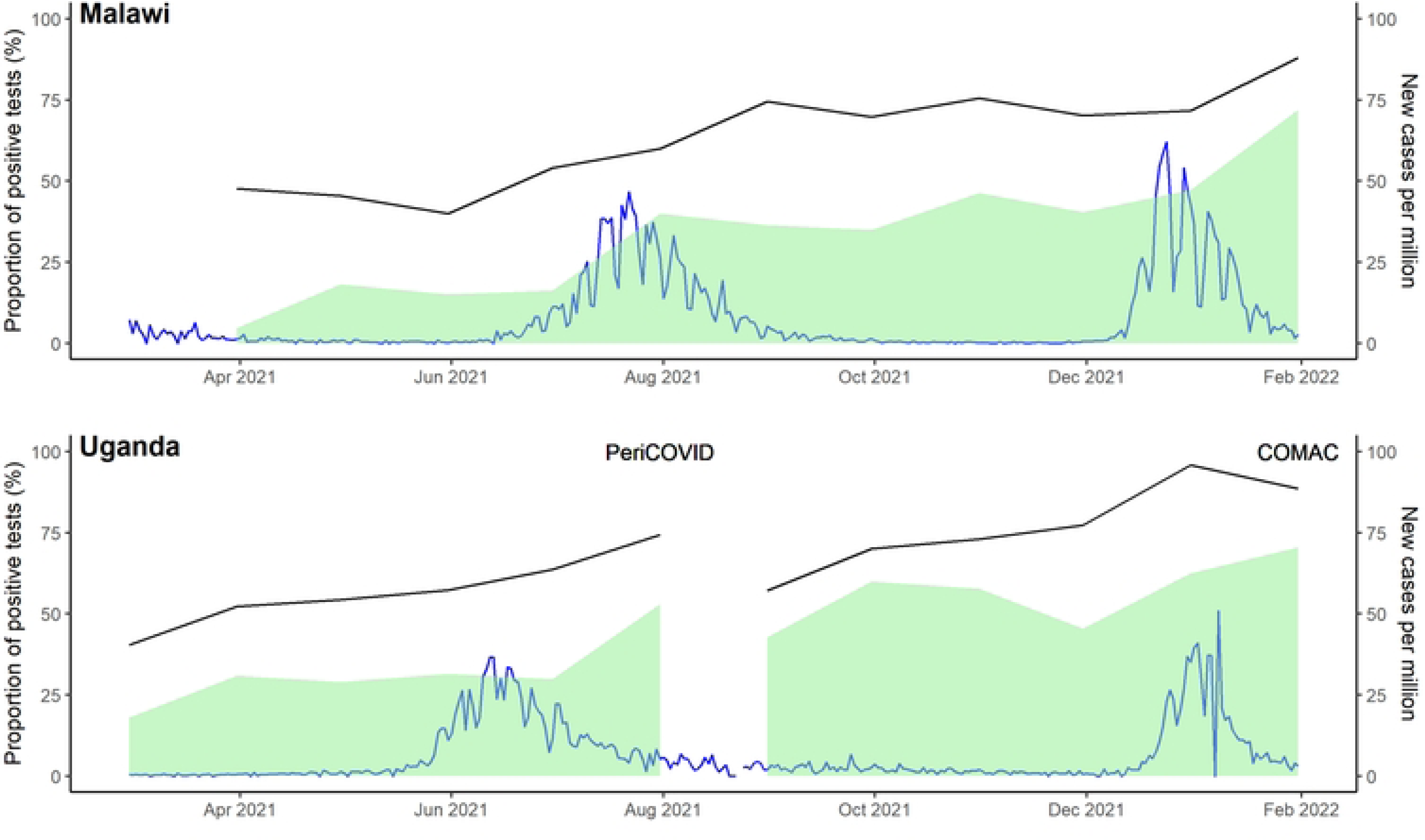
Monthly sero-positivity by country in pregnant women. Black line is the monthly proportion of results that were positive. Shaded in green is the proportion of Wantai positive samples that were also positive for anti-spike IgG in the Euroimmune assay. Blue line is the number of new cases per million in Malawi and Uganda, taken from Our World in Data.

**Table 3.**
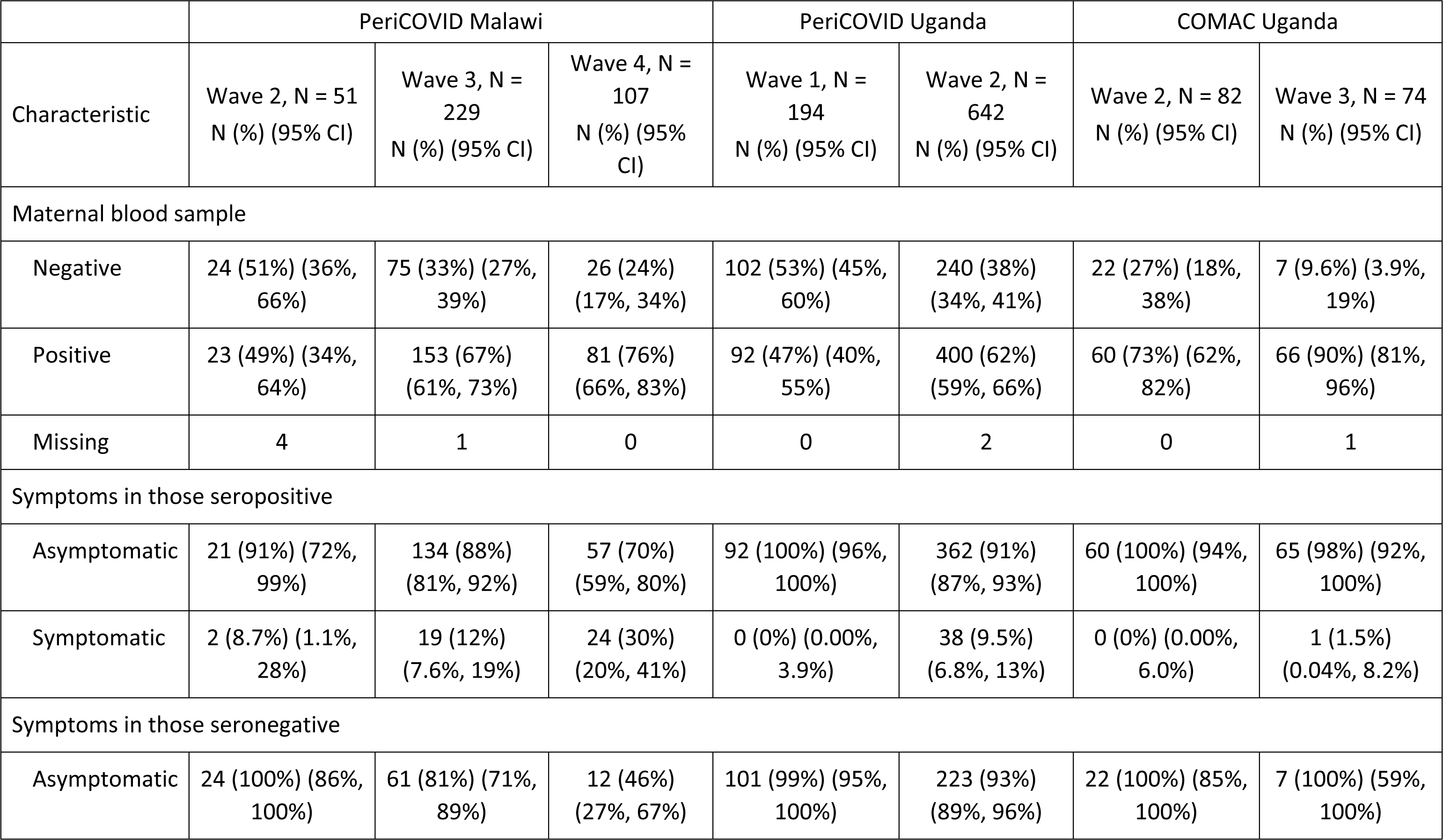
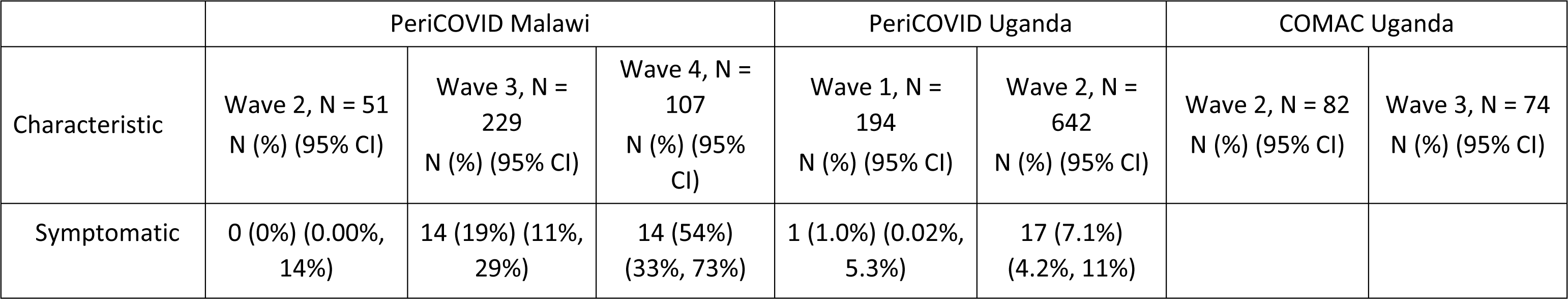
Maternal results.

|  | PeriCOVID Malawi |  |  | PeriCOVID Uganda |  | COMAC Uganda |  |
| --- | --- | --- | --- | --- | --- | --- | --- |
| Characteristic | Wave 2, N = 51<br>N (%) (95% CI) | Wave 3, N = 229<br>N (%) (95% CI) | Wave 4, N = 107<br>N (%) (95% CI) | Wave 1, N = 194<br>N (%) (95% CI) | Wave 2, N = 642<br>N (%) (95% CI) | Wave 2, N = 82<br>N (%) (95% CI) | Wave 3, N = 74<br>N (%) (95% CI) |
| Maternal blood sample |  |  |  |  |  |  |  |
| Negative | 24 (51%) (36%, 66%) | 75 (33%) (27%, 39%) | 26 (24%) (17%, 34%) | 102 (53%) (45%, 60%) | 240 (38%) (34%, 41%) | 22 (27%) (18%, 38%) | 7 (9.6%) (3.9%, 19%) |
| Positive | 23 (49%) (34%, 64%) | 153 (67%) (61%, 73%) | 81 (76%) (66%, 83%) | 92 (47%) (40%, 55%) | 400 (62%) (59%, 66%) | 60 (73%) (62%, 82%) | 66 (90%) (81%, 96%) |
| Missing | 4 | 1 | 0 | 0 | 2 | 0 | 1 |
| Symptoms in those seropositive |  |  |  |  |  |  |  |
| Asymptomatic | 21 (91%) (72%, 99%) | 134 (88%) (81%, 92%) | 57 (70%) (59%, 80%) | 92 (100%) (96%, 100%) | 362 (91%) (87%, 93%) | 60 (100%) (94%, 100%) | 65 (98%) (92%, 100%) |
| Symptomatic | 2 (8.7%) (1.1%, 28%) | 19 (12%) (7.6%, 19%) | 24 (30%) (20%, 41%) | 0 (0%) (0.00%, 3.9%) | 38 (9.5%) (6.8%, 13%) | 0 (0%) (0.00%, 6.0%) | 1 (1.5%) (0.04%, 8.2%) |
| Symptoms in those seronegative |  |  |  |  |  |  |  |
| Asymptomatic | 24 (100%) (86%, 100%) | 61 (81%) (71%, 89%) | 12 (46%) (27%, 67%) | 101 (99%) (95%, 100%) | 223 (93%) (89%, 96%) | 22 (100%) (85%, 100%) | 7 (100%) (59%, 100%) |
| Characteristic | Wave 2, N = 51<br>N (%) (95% CI) | Wave 3, N = 229<br>N (%) (95% CI) | Wave 4, N = 107<br>N (%) (95% CI) | Wave 1, N = 194<br>N (%) (95% CI) | Wave 2, N = 642<br>N (%) (95% CI) | Wave 2, N = 82<br>N (%) (95% CI) | Wave 3, N = 74<br>N (%) (95% CI) |
| Symptomatic | 0 (0%) (0.00%, 14%) | 14 (19%) (11%, 29%) | 14 (54%) (33%, 73%) | 1 (1.0%) (0.02%, 5.3%) | 17 (7.1%) (4.2%, 11%) |  |  |

**Table 4.** Cord blood serology.

|  | PeriCOVID Malawi |  |  | PeriCOVID Uganda |  | COMAC Uganda |  |
| --- | --- | --- | --- | --- | --- | --- | --- |
| Characteristic | Wave 2, N = 52<br>N (%) (95% CI) | Wave 3, N = 230<br>N (%) (95% CI) | Wave 4, N = 111<br>N (%) (95% CI) | Wave 1, N = 195<br>N (%) (95% CI) | Wave 2, N = 642<br>N (%) (95% CI) | Wave 2, N = 82<br>N (%) (95% CI) | Wave 3, N = 74<br>N (%) (95% CI) |
| Concordance of maternal and cord blood samples |  |  |  |  |  |  |  |
| Concordant | 35 (80%) (65%, 90%) | 194 (92%) (88%, 96%) | 101 (96%) (91%, 99%) | 79 (82%) (73%, 89%) | 271 (72%) (67%, 76%) | 81 (99%) (93%, 100%) | 68 (96%) (88%, 99%) |
| Discordant | 9 (20%) (9.8%, 35%) | 16 (7.6%) (4.4%, 12%) | 4 (3.8%) (1.0%, 9.5%) | 17 (18%) (11%, 27%) | 106 (28%) (24%, 33%) | 1 (1.2%) (0.03%, 6.6%) | 3 (4.2%) (0.88%, 12%) |
| Missing | 8 | 20 | 6 | 99 | 265 | 0 | 3 |
| Cord blood results with positive maternal blood result |  |  |  |  |  |  |  |
| Negative | 8 (40%) (19%, 64%) | 14 (10%) (5.7%, 17%) | 3 (3.8%) (0.78%, 11%) | 8 (18%) (8.2%, 33%) | 38 (16%) (12%, 21%) | 0 (0%) (0.00%, 6.0%) | 3 (4.7%) (0.98%, 13%) |
| Positive | 12 (60%) (36%, 81%) | 123 (90%) (83%, 94%) | 77 (96%) (89%, 99%) | 36 (82%) (67%, 92%) | 198 (84%) (79%, 88%) | 60 (100%) (94%, 100%) | 61 (95%) (87%, 99%) |
| Missing | 3 | 15 | 3 | 48 | 165 | 0 | 2 |
| Cord blood result with negative maternal blood result |  |  |  |  |  |  |  |
| Negative | 23 (96%) (79%, 100%) | 71 (97%) (90%, 100%) | 24 (96%) (80%, 100%) | 43 (83%) (70%, 92%) | 73 (52%) (43%, 60%) | 21 (95%) (77%, 100%) | 7 (100%) (59%, 100%) |
| Characteristic | Wave 2, N = 52<br>N (%) (95% CI) | Wave 3, N = 230<br>N (%) (95% CI) | Wave 4, N = 111<br>N (%) (95% CI) | Wave 1, N = 195<br>N (%) (95% CI) | Wave 2, N = 642<br>N (%) (95% CI) | Wave 2, N = 82<br>N (%) (95% CI) | Wave 3, N = 74<br>N (%) (95% CI) |
| Positive | 1 (4.2%)<br>(0.11%, 21%) | 2 (2.7%)<br>(0.33%, 9.5%) | 1 (4.0%)<br>(0.10%, 20%) | 8 (15%) (6.9%, 28%) | 68 (48%) (40%, 57%) | 1 (4.5%)<br>(0.12%, 23%) | 0 (0%) (0.00%, 41%) |
| Missing | 1 | 4 | 3 | 52* | 98 |  |  |

\* 1 cord blood sample in PeriCOVID Uganda was insufficient for analysis

### SARS-CoV-2 infection in pregnancy and key adverse pregnancy and neonatal outcomes

1220/1224 mothers had serology results at enrolment available for analysis and were included in the analysis of Sars-CoV-2 infection and pregnancy outcomes. 79/1220 mothers experienced at least one adverse pregnancy outcome (maternal death N=4, abortion N=4, premature labour N=52, and stillbirth N=26) and there were 46 infant deaths (Supp Table 4 and 5). Of the 79 adverse pregnancy outcomes, 34 (43%) mothers were sero-negative, and 45 (57%) mothers were sero-positive, compared to 435 (38%) mothers who were sero-negative and 706 (62%) mothers who were sero-positive with no adverse pregnancy outcomes. There was no difference in pregnancy outcomes due to sero-positive SARS-CoV-2 status (Table 5). Due to the small numbers of outcomes, the impact of Sars-CoV-2 infection on maternal death could not be modelled. There was also no difference in outcomes due to SARS-CoV-2 infection with and without symptoms in the 14 days (periCOVID Uganda and Malawi) −28 days (COMAC) prior to enrolment (Supp Table 6).

**Table 5.** Impact of infection (seropositivity) on key pregnancy and neonatal outcomes in Pericovid Malawi and Pericovid Uganda.

|  | Number included in model | Number of events | Relative Risk | 95% Confidence Interval |
| --- | --- | --- | --- | --- |
| **Maternal death** |  |  |  |  |
| Sero-Negative |  | 1 | — | — |
| Sero-Positive |  | 3 | — | — |
| **Infant death** |  |  |  |  |
| Sero-Negative | 471 | 20 | — | — |
| Sero-Positive | 752 | 26 | 0.81 | 0.46, 1.46 |
| **Premature labour** |  |  |  |  |
| Sero-Negative | 476 | 20 | — | — |
| Sero-Positive | 758 | 32 | 0.97 | 0.57, 1.71 |
| **Still birth** |  |  |  |  |
| Sero-Negative | 473 | 15 | — | — |
| Sero-Positive | 752 | 11 | 0.48 | 0.22, 1.03 |
| **Abortion** |  |  |  |  |
| Sero-Negative |  | 0 | — | — |
| Sero-Positive |  | 4 | — | — |
| **Combined adverse pregnancy outcome** |  |  |  |  |
| Sero-Negative | 463 | 34 | — | — |
| Sero-Positive | 742 | 45 | 0.81 | 0.53, 1.26 |
| **Low birth weight** |  |  |  |  |
| Sero-Negative | 474 | 22 | — | — |
| Sero-Positive | 753 | 39 | 0.98 | 0.60, 1.65 |
| **NICU admission** |  |  |  |  |
| Sero-Negative | 471 | 63 | — | — |
| Sero-Positive | 753 | 107 | 0.96 | 0.73, 1.28 |
| **Combined adverse neonatal outcome** |  |  |  |  |
| Sero-Negative | 476 | 77 | — | — |
| Sero-Positive | 758 | 120 | 0.92 | 0.71, 1.19 |

Models are adjusted for country

There were 197 adverse infant outcomes (at least one of: infant/neonatal death, prematurity, low birth weight, NICU admission and birth asphyxia). There were 46 infant deaths, 26 (57%) from sero-positive and 20 (43%) from sero-negative women (Supp Table 5). There was no difference in risk of infant death due to SARS-CoV-2 serology status in the mother (Table 5). 77 (39%) infants with adverse outcomes were born to women who were sero-negative and 120 (61%) sero-positive. The relative risk was 0.92 (95% CI 0.71, 1.19), providing no evidence of a difference in the risk of at least one adverse neonatal outcome due to serological status of the mother. There was also no evidence of a difference in neonatal outcome due to positive serological status with and without symptoms in the mother (Supp Tables 4 and 5).

### Placental transfer of SARS-CoV-2 antibodies in those with prior infection and/or vaccination

In PeriCOVID Uganda, 208/503 mother-infant pairs had anti-S IgG results available for analysis, corresponding to 27 and 181 during the first and second waves respectively. There was no difference between the maternal and cord blood anti-S IgG GMCs during the first wave. The GMR in the second wave was 1.7 (1.3, 2.3), indicating that anti-S IgG was higher in the cord blood than the maternal blood at enrolment. In comparison, in COMAC Uganda the GMR indicated no difference between the maternal and cord blood anti-S IgG for the 60 mother-infant pairs enrolled during the second wave, or for the 59 enrolled during the third wave (Supp Table 7). The rate of placental transfer of anti-S IgG is plotted in Fig 3.

**Fig 3.**
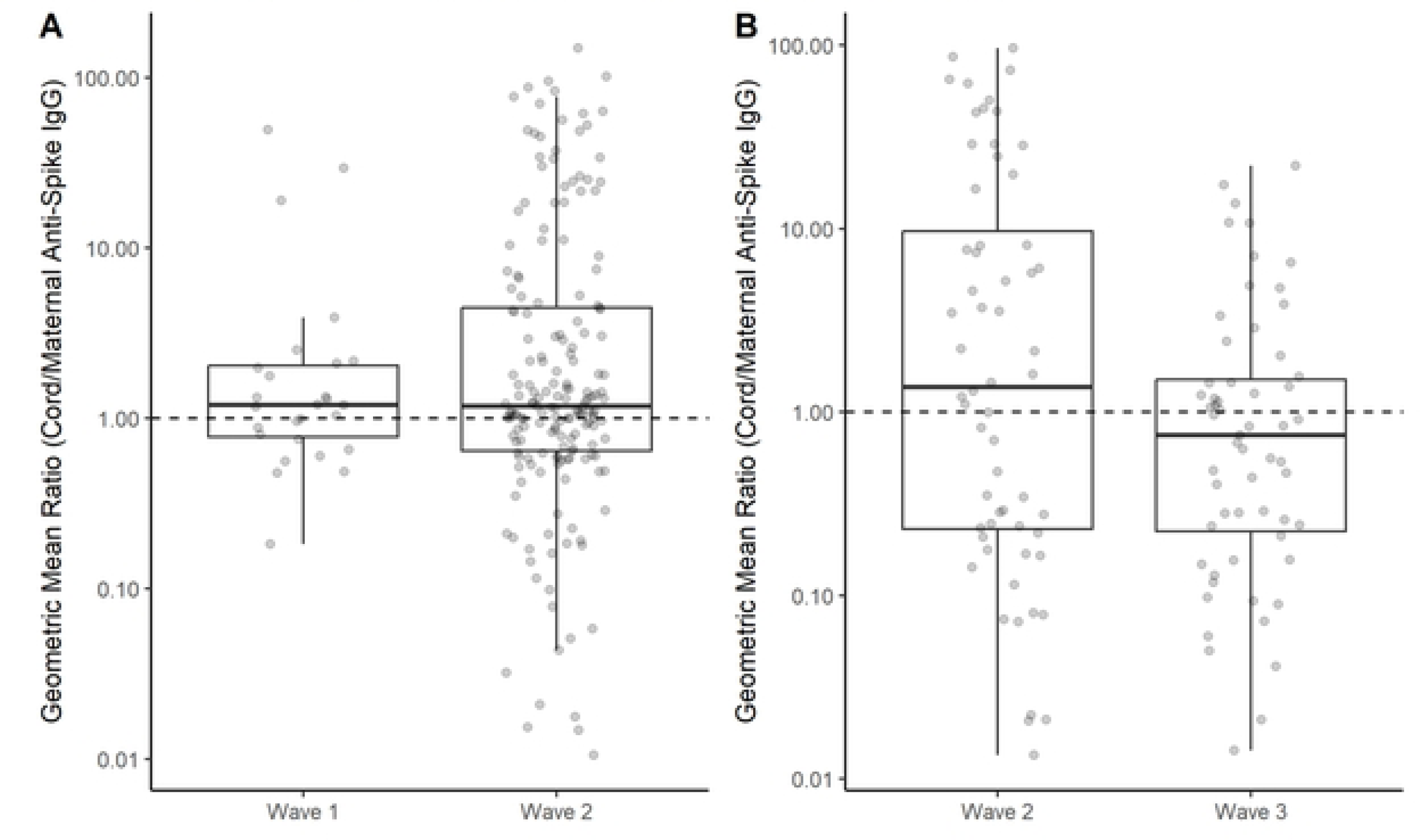
Placental transfer of anti-s IgG in A) PeriCOVID Uganda and B) COMAC Uganda. Geometric mean ratios (GMRs) of anti-spike IgG for each mother-infant pair with results available for analysis on the Euroimmune anti-spike IgG assay. Boxplots of the GMRs show no evidence of a difference in placental transfer in different waves of the pandemic.

There were 39 mother-infant pairs enrolled in PeriCOVID Uganda during the first wave with anti-NCP results, and 194 during the second wave. For both waves, there was no evidence of a difference in the maternal and cord blood anti-NCP IgG results. In COMAC Uganda, 55 and 43 mother-infant pairs enrolled during the second and third waves, respectively, had anti-NCP IgG results. The GMR in the second wave was 1.8 (1.1, 2.9), and in the third wave was 2.7 (1.7, 4.4), indicating that anti-NCP IgG was higher in cord blood samples than in the maternal blood (Supp Table 7). The rate of placental transfer of anti-NCP IgG is plotted in Fig 3. There was no clear evidence of a difference in placental transfer of both anti-S and anti-NCP IgG during different waves (Fig 3 and Fig S1).

### Number of vaccinated women during pregnancy

A total of 29 participants across all sites reported prior vaccination to SARS-CoV-2: 7 from Malawi, 1 from periCOVID Uganda and 21 from COMAC. Monthly numbers of positive results in COMAC Uganda by vaccination status can be seen in Fig S2.

## Discussion

This study describes the increasing prevalence of SARS CoV-2 infection across 2 countries and 5 hospital sites in East and southern Africa, and across several COVID-19 waves. This increase in prevalence coincided with waves of delta and omicron infection within the countries, respectively. Our data indicates that the majority of cases were asymptomatic and adds to existing evidence that suggests under-reporting of infection if based solely on confirmed cases by PCR(4, 5, 21).

The high prevalence of poor maternal and child health outcomes in sub-Saharan Africa, combined with the known impact of SARS-CoV-2 illness in pregnancy from existing studies outside of Africa (22–26), means that we need to better understand the direct effects of exposure to COVID-19 in pregnancy and outcome for the pregnant woman and her infant. The INTERCOVID study (27), which included 2 sites in West Africa (Nigeria and Ghana) showed that infection in pregnancy was associated with increased maternal and neonatal morbidity and mortality. The AFREHEALTH study of 1315 hospitalized pregnant and non-pregnant women with and without SARS-CoV-2 revealed an increased risk of ICU admission and in-hospital death amongst pregnant women with COVID-19 (28). Though we are unable to assess outcomes for symptomatic SARS-CoV-2 illness in our cohort due to low numbers our study does highlight that asymptomatic infection does not appear to be associated with death in the mother, or with worse neonatal outcomes in the first month of life. This is reassuring to parents and health care providers. Furthermore, in our study placental transfer of IgG increased during each subsequent wave. Previous studies earlier in the pandemic suggested reduced placental transfer of IgG in women with a positive SARS-CoV-2 RT-PCR (29, 30). More research is needed to better understand placental transfer with different SARS-CoV-2 variants.

### Limitations

Our study is limited by differences in methodology across sites, with sampling performed at different time points in pregnancy, despite efforts to adhere to the UNITY protocol. The uniform collection of cord blood across all sites however enables comparison across sites and strengthens our results. In PeriCOVID Uganda, a strict period of lockdown over the summer of 2021 with a ban on public transport made it challenging for participants to attend hospital for delivery, leading to a lower number of cord blood samples than anticipated. Calls to participants by healthcare visitors were increased to ensure that women were aware that study staff were still working and could care for them during their delivery. We report our primary outcomes using the Wantai assay, but for placental antibody transfer, we report IgG using Euroimmune results. However, we identified cross-reactivity of antibodies against *Plasmodium falciparum* or other common cold coronaviruses (CCCs) as has been reported elsewhere in East Africa (21), meaning these results should be reviewed with caution. We had initially planned to use the Euroimmune anti-NCP assay to differentiate between infection and vaccination. However, the specificity of the assay precluded its use for this purpose. The Euroimmune anti-NCP assay has a lower sensitivity than other assays (31). Several studies have shown a low anti-NCP positivity after mild infections (32). As the pandemic progressed the chance of repeat SARS-CoV-2 infection increased, though these infections were generally milder (32). Anti-NCP antibodies in some studies have remained negative in individuals who were vaccinated against SARS-CoV-2 and who had an rt-PCR confirmed illness (32). A lower anti-NCP seropositivity later in the pandemic may therefore represent assay sensitivity and a lower anti-NCP immune response following mild or asymptomatic infection.

## Conclusion

Data from Uganda and Malawi showed a seroprevalence of SARS-CoV-2 higher than the cases figures identified by other sources. In future pandemics and outbreaks, seroprevalence studies may be a more accurate measure in assessing the true prevalence of infection.

## Data Availability

The data dictionary for results presented in this study are available from St George's University, Figshare (https://sgul.figshare.com/)

## Acknowledgments

The periCOVID Consortium consists of:

Esperança Sevene^1,2^, Sónia Maculuve^1^, Anifa Valá^1^, Angela Koech^3^, Geoffrey Omuse^4^, Marleen Temmerman^3^

1. Manhiça Health Research Center, Manhiça, Mozambique
2. Department of Physiological Science, Faculty of Medicine, Eduardo Mondlane University, Maputo, Mozambique
3. Centre of Excellence in Women and Child Health, Aga Khan University, Kenya
4. Department of Pathology, Aga Khan University, Kenya

We would also like to thank all study and laboratory staff who enabled recruitment, follow-up, and study testing.

## Supplementary Information

**S1 Fig.**
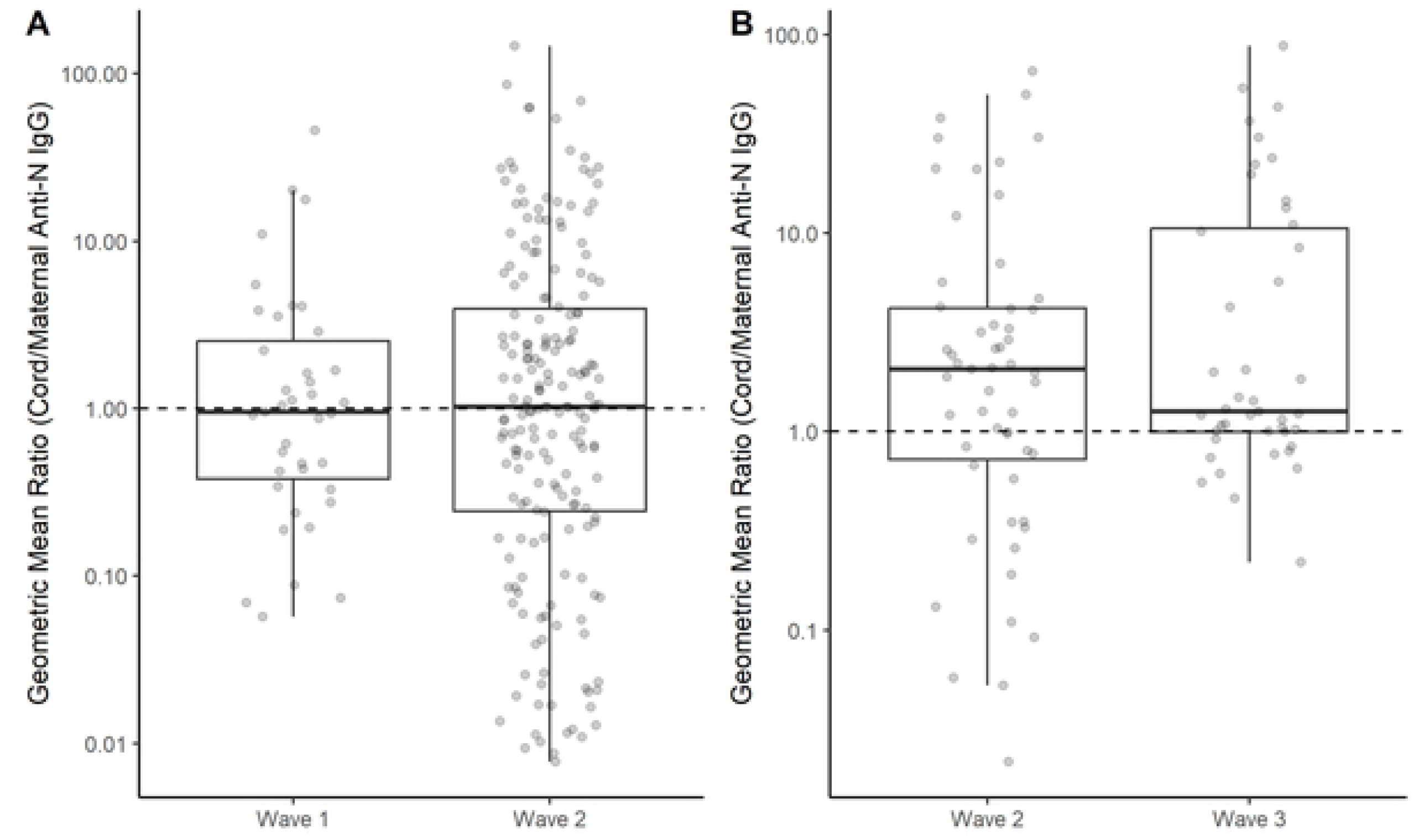
Placental transfer of anti-n IgG in A) PeriCOVID Uganda and B) COMAC Uganda. Geometric mean ratios (GMRs) of anti-nucelocapsid IgG for each mother-infant pair with results available for analysis on the Euroimmune anti-nucleocapsid IgG assay. Boxplots of the GMRs show no evidence of a difference in placental transfer in different waves of the pandemic

**S2 Fig.**
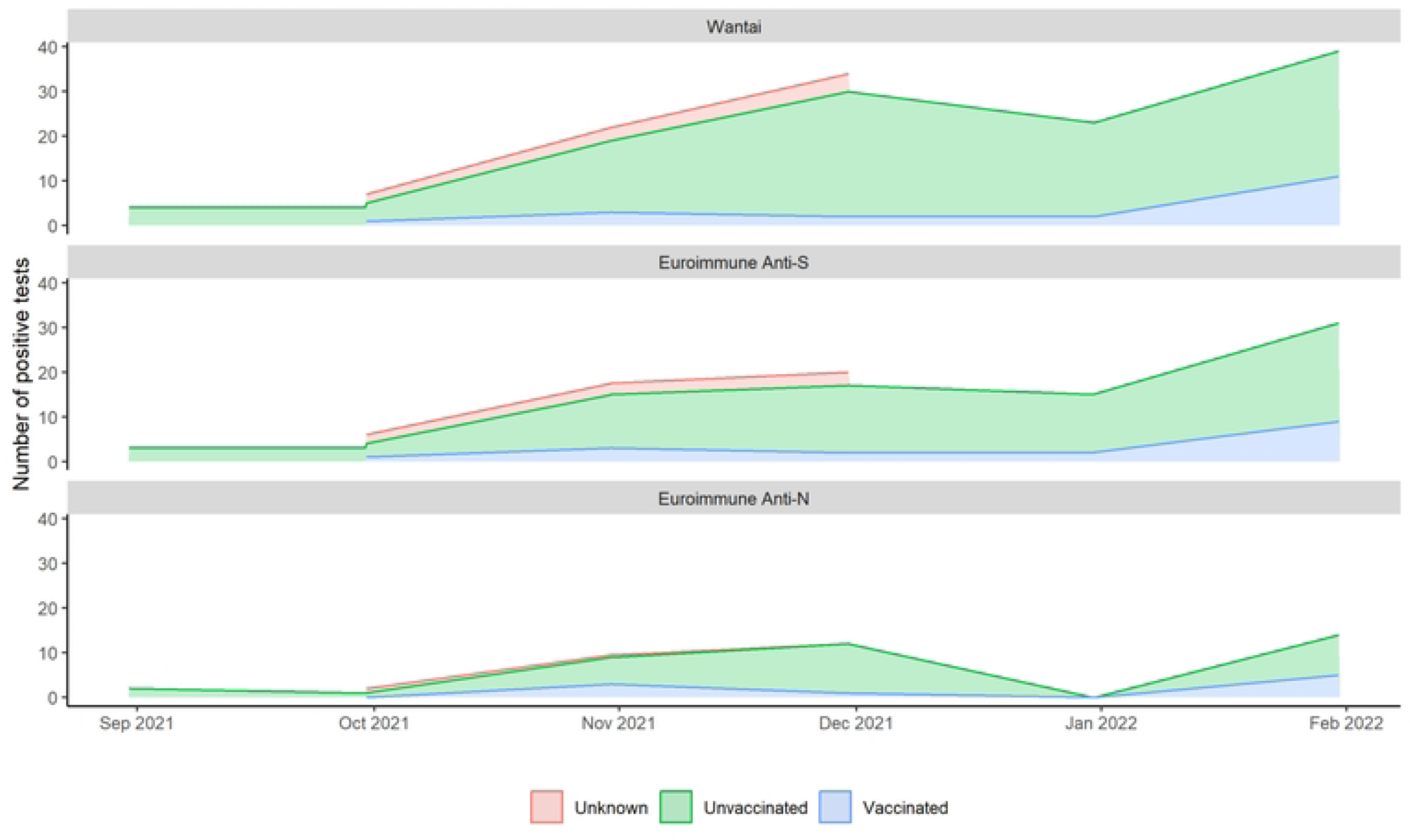
Monthly number of positives in COMAC Uganda, by vaccination status. The monthly number of women in COMAC Uganda who were seropositive at enrolment on each of the Wantai (top panel), Euroimmune anti-S (middle panel) and Euroimmune anti-N (bottom panel) assays, coloured by vaccination status. Green shows those who were unvaccinated at enrolment, blue shows the small proportion who were vaccinated, and red shows those whose vaccination status was unknown at enrolment. The majority were unvaccinated.

**S1 Table.** Wantai Assay Specificity.

|  | Wantai positive (n) | Wantai negative (n) | Specificity (%) |
| --- | --- | --- | --- |
| 2019 samples, malaria positive (n=74) | 1 | 73 | 98.65 |
| 2019 samples, malaria negative (n=100) | 1 | 99 | 99 |

**S2 Table.** Dates used to define COVID-19 waves in Malawi and Uganda.

| Country | Wave 1 | Wave 2 | Wave 3 | Wave 4 |
| --- | --- | --- | --- | --- |
| Uganda | 26/03/2020 –<br>21/03/2021 | 22/03/2021 –<br>30/11/2021 | 01/12/2021 –<br>30/04/2022 |  |
| Malawi | 07/04/2020 –<br>16/11/2020 | 17/11/2020 –<br>15/05/2021 | 16/05/2021 –<br>11/11/2021 | 12/11/2021 –<br>20/04/2022 |

**S3 Table.** Maternal PCR tests by study site and wave.

|  | PeriCOVID Malawi |  |  |  | PeriCOVID Uganda |  |  |
| --- | --- | --- | --- | --- | --- | --- | --- |
| Characteristic | Overall, N = 73<br>(95% CI)<sup>12</sup> | Wave 2, N = 2<br>(95% CI)<sup>12</sup> | Wave 3, N = 31<br>(95% CI)<sup>12</sup> | Wave 4, N = 40<br>(95% CI)<sup>12</sup> | Overall, N = 836<br>(95% CI)<sup>12</sup> | Wave 1, N = 194<br>(95% CI)<sup>12</sup> | Wave 2, N = 642<br>(95% CI)<sup>12</sup> |
| PCR result |  |  |  |  |  |  |  |
| Negative | 24 (33%) (22%, 45%) | 1 (50%) (1.3%, 99%) | 11 (35%) (19%, 55%) | 12 (30%) (17%, 47%) | 817 (98%) (96%, 99%) | 194 (100%) (98%, 100%) | 623 (97%) (95%, 98%) |
| Positive | 49 (67%) (55%, 78%) | 1 (50%) (1.3%, 99%) | 20 (65%) (45%, 81%) | 28 (70%) (53%, 83%) | 19 (2.3%) (1.4%, 3.5%) | 0 (0%) (0.00%, 1.9%) | 19 (3.0%) (1.8%, 4.6%) |
| Symptoms in those with positive PCR |  |  |  |  |  |  |  |
| Asymptomatic | 2 (4.1%) (0.50%, 14%) | 0 (0%) (0.00%, 98%) | 0 (0%) (0.00%, 17%) | 2 (7.1%) (0.88%, 24%) | 13 (68%) (43%, 87%) | NA | 13 (68%) (43%, 87%) |
| Symptomatic | 47 (96%) (86%, 100%) | 1 (100%) (2.5%, 100%) | 20 (100%) (83%, 100%) | 26 (93%) (76%, 99%) | 6 (32%) (13%, 57%) | NA | 6 (32%) (13%, 57%) |

Symptoms are those defined as probable infection according to WHO criteria (18)

**S4 Table.** Adverse pregnancy outcomes in mothers enrolled in PeriCOVID Malawi and PeriCOVID Uganda.

|  | **All Enrolled** | **Wantai Overall** | **Wantai Serostatus** |  |
| --- | --- | --- | --- | --- |
| **Characteristic** | **N = 1,224<sup>1</sup>** | **N = 1,220<sup>1</sup>** | **Negative, N = 469<sup>2</sup>** | **Positive, N = 751<sup>2</sup>** |
| Maternal death | 4 (0.3%) | 4 (0.3%) | 1 (25%) | 3 (75%) |
| Unknown | 27 | 25 | 9 | 16 |
| Abortion | 4 (0.3%) | 4 (0.3%) | 0 (0%) | 4 (100%) |
| Unknown | 25 | 24 | 9 | 15 |
| Premature labour | 52 (4.3%) | 52 (4.3%) | 20 (38%) | 32 (62%) |
| Unknown | 1 |  |  |  |
| Stillbirth | 26 (2.1%) | 26 (2.1%) | 15 (58%) | 11 (42%) |
| At least one adverse pregnancy outcome | 79 (6.5%) | 79 (6.5%) | 34 (43%) | 45 (57%) |
| No adverse pregnancy outcome | 1,145 (94%) | 1,141 (94%) | 435 (38%) | 706 (62%) |
| Livebirth | 1,198 (98%) | 1,194 (98%) | 454 (38%) | 740 (62%) |

<sup>1</sup> Column percentages are presented for the overall number of women experiencing each outcome
<sup>2</sup> Row percentages are presented for the number of seropositive and seronegative women experiencing each outcome

**S5 Table.** Adverse neonatal outcomes in infants born in PeriCOVID Malawi and PeriCOVID Uganda.

|  | **All infants** | **Wantai Overall** | **Maternal Wantai Serostatus** |  |
| --- | --- | --- | --- | --- |
| **Characteristic** | **N = 1,230<sup>1</sup>** | **N = 1,227<sup>1</sup>** | **Negative, N = 474<sup>2</sup>** | **Positive, N = 753<sup>2</sup>** |
| Infant death | 46 (3.8%) | 46 (3.8%) | 20 (43%) | 26 (57%) |
| Unknown | 5 | 4 | 3 | 1 |
| Prematurity | 58 (4.7%) | 58 (4.7%) | 22 (38%) | 36 (62%) |
| Unknown | 1 |  |  |  |
| Low birth weight | 61 (5.0%) | 61 (5.0%) | 22 (36%) | 39 (64%) |
| Unknown | 1 |  |  |  |
| Admitted to NICU | 170 (14%) | 170 (14%) | 63 (37%) | 107 (63%) |
| Unknown | 4 | 3 | 3 | 0 |
| Birth asphyxia | 53 (4.3%) | 53 (4.3%) | 20 (38%) | 33 (62%) |
| Unknown | 1 |  |  |  |
| At least one adverse neonatal outcome | 197 (16%) | 197 (16%) | 77 (39%) | 120 (61%) |
| No adverse neonatal/infant outcome | 1,033 (84%) | 1,030 (84%) | 397 (39%) | 633 (61%) |

<sup>1</sup> Column percentages are presented for the overall number of infants experiencing each outcome
<sup>2</sup> Row percentages are presented for the number of infants born to seropositive and seronegative women with each outcome

**S6 Table.** Impact of infection (seropositive and WHO probable) on key pregnancy and neonatal outcomes.

|  | Number included in model | Number of events | Relative Risk | 95% Confidence Interval |
| --- | --- | --- | --- | --- |
| **Maternal death** |  |  |  |  |
| Infection status |  | 4 |  |  |
| sero-negative |  | 1 | — | — |
| sero-positive and not WHO probable |  | 0 | — | — |
| sero-positive and WHO probable |  | 3 | — | — |
| **Infant death** |  |  |  |  |
| Infection status | 1,223 | 46 |  |  |
| sero-negative | 471 | 20 | — | — |
| sero-positive and not WHO probable | 667 | 24 | 0.85 | 0.47, 1.53 |
| sero-positive and WHO probable | 85 | 2 | — | — |
| **Premature labour** |  |  |  |  |
| Infection status | 1,234 | 52 |  |  |
| Sero- negative | 476 | 20 | — | — |
| Sero- positive and not WHO probable | 672 | 27 | 0.94 | 0.53, 1.67 |
| Sero- positive and WHO probable | 86 | 5 | 1.24 | 0.42, 3.00 |
| **Still birth** |  |  |  |  |
| Infection status | 1,225 | 26 |  |  |
| Sero- negative | 473 | 15 | — | — |
| Sero- positive and not WHO probable | 667 | 10 | 0.48 | 0.21, 1.05 |
| Sero- positive and WHO probable | 85 | 1 | — | — |
| **Abortion** |  |  |  |  |
| Infection status |  | 4 |  |  |
| Sero- negative |  | 0 | — | — |
| Sero- positive and not WHO probable |  | 4 | — | — |
| Sero- positive and WHO probable |  | 0 | — | — |
| **Combined adverse pregnancy outcome** |  |  |  |  |
| Infection status | 1,205 | 79 |  |  |
| Sero- negative | 463 | 34 | — | — |
| Sero- positive and not WHO probable | 659 | 36 | 0.74 | 0.47, 1.16 |
| Sero- positive and WHO probable | 83 | 9 | 1.39 | 0.64, 2.68 |
| **Low birth weight** |  |  |  |  |
| Infection status | 1,227 | 61 |  |  |
| Sero- negative | 474 | 22 | — | — |
| Sero- positive and not WHO probable | 668 | 33 | 0.99 | 0.59, 1.67 |
| Sero- positive and WHO probable | 85 | 6 | 0.98 | 0.37, 2.17 |
| **NICU admission** |  |  |  |  |
| Infection status | 1,224 | 170 |  |  |
| Sero- negative | 471 | 63 | — | — |
| Sero- positive and not WHO probable | 668 | 90 | 0.95 | 0.72, 1.28 |
| Sero- positive and WHO probable | 85 | 17 | 1.03 | 0.62, 1.60 |
| **Combined adverse neonatal outcome** |  |  |  |  |
| Infection status | 1,234 | 197 |  |  |
| Sero- negative | 476 | 77 | — | — |
| Sero- positive and not WHO probable | 672 | 102 | 0.91 | 0.70, 1.19 |
| Sero- positive and WHO probable | 86 | 18 | 0.98 | 0.60, 1.50 |

Models are adjusted for country

**S7 Table.** Placental Transfer.

Models are adjusted for country
|  |  | PeriCOVID Uganda |  | COMAC Uganda |  |
| --- | --- | --- | --- | --- | --- |
|  |  | Wave 1 | Wave 2 | Wave 2 | Wave 3 |
| Anti-S | Infant, GMC (95% CI) | 78.8 (46.4, 133.7) [N = 27] | 127.6 (100.6, 161.8) [N = 181] | 152 (100.3, 230.3) [N = 60] | 65.2 (47.1, 90.4) [N = 59] |
|  | Mother, GMC (95% CI) | 51.7 (34.6, 77.3) [N = 27] | 74.7 (59.3, 94.2) [N = 181] | 96.1 (65.9, 140.1) [N = 60] | 98.5 (68.6, 141.5) [N = 59] |
|  | Placental transfer, GMR (95% CI) | 1.5 (0.9, 2.5) [N = 27] | 1.7 (1.3, 2.3) [N = 181] | 1.6 (0.8, 3) [N = 60] | 0.7 (0.4, 1) [N = 59] |
| Anti-N | Infant, GMC (95% CI) | 57.1 (38, 85.8) [N = 39] | 133.7 (106.5, 167.8) [N = 194] | 155.9 (102.1, 237.9) [N = 55] | 172 (103, 287.2) [N = 43] |
|  | Mother, GMC (95% CI) | 57.6 (41.5, 79.9) [N = 39] | 146.2 (114.4, 186.8) [N = 194] | 87.6 (54.9, 139.8) [N = 55] | 62.9 (35.5, 111.4) [N = 43] |
|  | Placental transfer, GMR (95% CI) | 1 (0.6, 1.6) [N = 39] | 0.9 (0.7, 1.3) [N = 194] | 1.8 (1.1, 2.9) [N = 55] | 2.7 (1.7, 4.4) [N = 43] |

